# Enabling Advanced Multi-Modal Neuroimaging Analysis within a Trusted Research Environment

**DOI:** 10.1101/2024.02.13.24302751

**Authors:** L Hotchkiss, E Squires, J E Gallacher, M Newbury, C Morris, RA Lyons, S Thompson

## Abstract

**Introduction:** Globally, 55 million individuals have dementia, with an increasing annual incident of 10 million. Enabling development of new multi-modal models can improve the current diagnostic pathways and potentially contribute to early diagnosis and treatment of dementia. Here, we report how multi-modal resources is achieved within the successful Trusted Research Environment (TRE) providing access to 60+ cohort datasets for dementia research, the Dementias Platform UK (DPUK).

**Objectives:** We aimed to identify the challenges of the storage, distribution and analysis of neuroimaging data and how we could implement a comprehensive infrastructure to deal with these. The problems we specifically aimed to address were how to: anonymise scans, store large amounts of data, standardise datasets to a common format, extract metadata, provision the data, and allow for analysis.

**Methods:** While, data within majority of existing research platforms are focused on a single aspect, DPUK data provides an enriched view of disease dynamic for dementia cohorts by providing access to linkable brain imaging and genomic data at the individual-level. We document various stages and capacities required for multi-modal neuroimaging analysis for dementia and conclude that achieving research ready assets to enable neuroimaging analysis for dementia from existing resources requires an engineered process to facilitate multiple aspects of curation, provisioning and large scale analysis.

**Results:** We developed an ingest pipeline for neuroimaging data to meet the requirements set out in the objectives. This involved standardising all datasets to the Brain Imaging Data Structure, defacing scans and anonymising data, using MinIO for data storage and extracting metadata from header information for data discovery and provisioning.

**Conclusion:** The neuroimaging ingest pipeline developed has allowed for the distribution of imaging datasets within DPUK which has facilitated multi-modal research on anonymised and standardised data. Our pipelines create research-ready datasets in a simplified way, reducing the time and effort of getting these datasets ready for data sharing and making the process easier for the data owners.

## Introduction

### Neuroimaging Analysis in Dementia Research

The World Health Organisation (WHO) estimates that 55.2 million people worldwide are living with dementia and forecasts that by 2050, this figure will rise to 139 million [1]. The total cost of care for people with dementia in the UK is £34.7 billion and this figure is predicted to rise to £94.1 billion by 2040 [2]. Therefore, research into biomarkers for diagnosis, prevention and treatment is imperative. Biomarkers can be defined as measurable substances in the body that act as reliable predictors and indicators of a disease process. Biomarkers make it possible to identify the disease before its clinical manifestation [3]. Over the last decade, advances have been made in developing biomarkers for Alzheimer’s disease (AD) using neuroimaging. This provides versatility in terms of understanding pathophysiological mechanisms. Magnetic resonance imaging (MRI) can be used to assess structural decline (e.g. loss in volume, cortical thinning), functional MRI (fMRI) can be used to assess functional decline (e.g. hyperactivity, altered network connectivity), diffusion MRI (dMRI) for white matter decline (e.g. diffusion anisotropy reduction, white matter pathology), whilst molecular imaging can be used to assess protein aggregation (e.g. amyloid and tau) using Positron Emission Tomography (PET)) [4].

To be able to analyse neuroimaging data, several pre-processing steps are usually required depending on the modality. Structural MRI scans, for example, require registration to align images to a standard space, noise reduction to reduce the amount of noise in the image from sources such as electrical interference or magnetic field variations, and segmentation to separate different structures or tissues for analysis. Functional MRI images often require additional processing steps, such as slice timing correction to align slices in time, motion correction to account for head movement, and spatial smoothing to reduce noise [5]. These pre-processing steps are computationally time-consuming and generate a substantial amount of data storage, which can be up to three times the original data size.

Advanced analytical techniques such as artificial intelligence (AI) are being used to identify biomarkers in dementia. AI uses a series of steps to identify, train, and test computer algorithms to identify a feature of interest. Some of these techniques have been applied to classify brain MRI or PET images, comparing dementia cases and healthy controls, and to distinguish different types or stages of dementia, cerebrovascular disease, and accelerated features of ageing [6]. Over the last few decades, the application of AI utilising neuroimaging data, has increased rapidly, largely due to the amplified availability and suitability of large neuroimaging datasets which facilitates the use of such techniques. However, one of the barriers for researchers using such data is the availability of access. This is because neuroimaging and health data of patients are extremely sensitive and of high volume which make it difficult for data sharing. However, it is imperative that such data sharing exists so that this research can be carried out.

### The Role of Trusted Research Environments (TRE’s) in Facilitating Access to Neuroimaging Data

A TRE is a secure computing environment that provides bona-fide researchers access to de-identified data to carry out approved projects. The secure environment ensures that data is accessed remotely within the environment, with data never leaving that environment. This governance arrangement allows data providers to be certain that anonymised data is only used for approved projects that meet the criteria of the onward sharing restrictions of that dataset.

Several platforms exist for holding and distributing neuroimaging data, including the Image and Data Archive (IDA) [7] and OpenNEURO [8]. However, neither of these are TRE’s as they are unable to provide a secure environment for accessing and using data. The IDA contains data from 144 studies and supports the de-identification of scans, but the distribution of these scans is primarily accomplished through direct downloads onto researchers’ personal desktops. A similar platform, OpenNEURO, also distributes through direct downloads but is additionally freely available instead of any agreements having to be signed. The way this is handled means that data owners may be reluctant to upload their data as these platforms don’t offer secure environments and tight controls on the data. It also means that users need to have adequate storage space to be able to download these large datasets which can often get to over 1TB in size. This means that they are restricted by the data that they can download and hinders the ability to be able to combine multiple large imaging datasets due to the size requirements.

Conversely, TRE’s are able to offer a suitable environment for the access and analysis of data without that data ever leaving the platform. The Birmingham-based TRE, PIONEER offers a secure environment for research to be carried out on a range of healthcare data, including imaging, and does anonymise data but is limited to data collected from University Hospitals Birmingham and the University of Birmingham [9]. There have been several TRE’s like this which have been set up to deal with limited data collected from specific places. However, more large-scale TRE’s exist, such as the Alzheimer’s Disease Data Initiative (ADDI), which includes data from multiple cohort studies [10]. However, ADDI is not focused on imaging data so has limited tools, processing and data available.

### The Challenges of using Advanced Techniques in Neuroimaging

#### Dataset Sizes

The application of AI in neuroimaging gives rise to several challenges due to small dataset sizes and high dimensionality. A typical MRI dataset contains a few hundred subject scans, with each sample containing thousands of features. This can result in overfitting and especially becomes a problem with extracting features from multiple modalities, creating even higher dimensionality [11]. Deep learning models, for example, have been shown to fit neuroimaging data very well, however, this often means that these models are not good at generalising. Consequently, this leads researchers to combine datasets to increase their sample sizes, however, this itself poses its challenges.

#### Combining Datasets

Combining datasets from across studies overcomes the problem of small sample sizes, but with this, comes the complication of variability between collection methods. Data, such as MRI scans from different study sites, are often acquired through varying methods and techniques. This is due to using different scanners and protocols, with no standardisation, which can yield different image qualities and characteristics [11]. Differences in participant demographics can also have an effect on using data across multiple cohorts. Although methods have been developed over recent years to overcome these differences between studies, there still exists the need for robust data harmonisation to combine datasets from multiple sites which relies on accurate acquisition data. Another problem with using data from multiple studies is that, often, studies will choose their own naming conventions and file structures to store their imaging data. This makes it difficult when trying to combine imaging data across studies since file structures and naming can vary, sometimes even making it difficult to determine the modality. Processing this data relies on well-structured directories with consistent naming to be able to perform analysis.

#### Storage Requirements

When large datasets are available, then a problem arises with storing that data and having enough computational power to be able to do research on it. Even smaller datasets can require large amounts of storage, for example, the Whitehall Imaging II sub-study (Filippini et al., 2014) contains 465 GB worth of data, from 800 participants. This easily gets into 1 TB worth of data once it has been pre-processed and analysed. With larger datasets, such as MEMENTO containing 2323 participants, or even the combination of multiple datasets, the requirements become higher, with the general expectation being that once a dataset is processed, then the data is likely to grow 2-3 fold, meaning that standard desktop computers are unable to cope with the sheer amount of data that is contained. In terms of storing this data in a TRE, these storage requirements further increase as multiple researchers use the datasets. This is due to researchers performing similar processing tasks and therefore creating large amounts of processed data which also has to be stored.

#### Computational Resources

This large amount of data also requires adequate computational power to be able to run the processing and analysis in a reasonable amount of time. FreeSurfer is a common toolkit for processing MRI scans, specifically structural [12]. However, on a standard desktop (4 cores, 16GB RAM), an individual subject can take up to around 20-30 hours to process, just to perform pre-processing and cortical reconstruction for a single structural scan. This is a long time to wait, especially with a dataset containing hundreds of subjects, with several varying types of scans. Taking into consideration a range of scans including T1-weighted, T2-weighted, Fluid-attenuated inversion recovery (FLAIR) and FMRI, it could take months for the different scans to be processed on a single participant’s data. One way that is typically used to reduce this time is to run analyses in parallel, however, most standard computers usually have only 4 or 8 cores. For a dataset containing 400 participants, with each subject taking 30 hours to process, without parallelisation, this could take 500 days. With parallelisation, utilising 4 cores, this is reduced to 125 days. When considering that datasets often contain several different modalities which, if longitudinal, will contain scans from several sessions. Then this time drastically increases and becomes a problem to do in a reasonable amount of time. FreeSurfer processes can take several hours, which makes the processing of hundreds of subjects, over multiple time periods, and with different modalities, a very time-inefficient process which often hinders researcher’s abilities to do the processing that they would want to do. When the processing is complete, then analysis of the data can also be computationally expensive due to the size of the data.

ML models often require high-performance power, especially due to the number of features generated by the processing. Robust ML models often also use several techniques such as cross-validation and optimisation, which are used to improve the performance of the models. However, these methods require repeated training of the model, which again significantly increases the time it takes to run analyses. Hence, the need for High-Performance Computing (HPC) clusters are often used to perform this processing as this makes more cores and memory available to researchers to significantly cut down the amount of processing time it takes.

## Methods

DPUK was established by the Medical research Council (MRC) in order to facilitate research into the treatment, cause and diagnosis of dementia. The DPUK data portal is a component infrastructure of DPUK which utilises the UK Secure eResearch Platform (UKSeRP) [13] to provide data discovery, access and analysis on a multitude of cohorts [14]. Over the years, with the growing amount of neuroimaging research and data collection, DPUK has acquired vast quantities of multi-modal imaging data. However, this has come with the need of finding new ways on how to handle, and perform analysis, on this large amount of data.

We examined the current UK neuroimaging data flows and identified seven levels of data facilitation via current platforms and TRE’s: 1) metadata tools, 2) data infrastructure, 3) anonymisation, 4) standardisation, 5) imaging data sharing, 6) multi-cohort access and 7) derived data, to assess the ability of existing systems for supporting neuroimaging data distribution and analysis. No platform or TRE has so far implemented all levels for neuroimaging, therefore, we aimed to implement each level of neuroimaging data facilitation.

### 1. Metadata

There are many platforms which provide metadata tools for researchers to find suitable imaging datasets to apply for and help facilitate that access by assisting in the application process. These are low-level platforms which don’t provide data themselves, but instead provide researchers with a useful resource to identify datasets. TRE’s have also developed similar metadata tools to showcase the data available within their environments, but these so far have focused on phenotypic data held within databases or csv files, with no current way of representing imaging or genomic metadata. We aimed to create metadata tools, by extracting metadata from the scans, to identify imaging datasets through categorising data by modalities. This information can be extracted either from DICOM headers or JSON

### 2. Data & Analysis Infrastructure

The problem with neuroimaging data in a TRE, because it is so large, is how to effectively store such large volumes of it and allow researchers to carry out processing and analysis. Therefore, we aimed to implement a Data Lake through MinIO [15], utilising object storage to store neuroimaging data. Object storage, unlike a typical hierarchy-based file storage system, stores data as unstructured objects that live in a flat address space and are identified by their unique address. This makes it ideal for storing imaging data in a way which is: scalable (infinitely to petabytes and beyond), efficient, and has customisable metadata. This means that large neuroimaging data can be stored at a reduced storage cost, so that the storage requirements can be met for researchers using this data and producing their own processed data. These objects are stored in buckets, where essentially, each neuroimaging dataset will be in its own bucket for users to access and use. Each object has customisable metadata and can be tagged to allow for easier retrieval and filtering of information. When it comes to provisioning data, data is tagged so that a filtered version of the bucket can be given to users based on certain filters such as a type of MRI scan, version, defaced or data from certain participants. This is useful as researchers can request specific types of scans from participants which means that they can then be given a filtered bucket with just the scans that they require. This is extremely beneficial for the point of view of a TRE, as data doesn’t need to be duplicated each time for each project as instead, they can just be given permissions to read certain objects in the bucket. This is useful in making both governance and storage more efficient since researchers are only given the data that they need. Because object storage can handle different types of data, AI/ML that needs to access data to create features means that different types of data matter, and the ability to be able to store, version and manage them from one place is truly important. Read/Write performance is also extremely important when accessing data due to the amount of data that these models need to use. MinIO has extremely fast read/write speeds which means that the performance of these models won’t be affected. Therefore, object storage provides an efficient way to provide users with data and a space to run their analyses and models on. MinIO also allows for tiering of data, so that datasets which are no longer used, or hardly used, can go in to deep storage, which essentially puts it on a storage drive which doesn’t require fast read/write speeds.

### 3. Anonymisation

Often, a requirement for sharing neuroimaging data is that scans should be anonymised by defacing structural scans, primarily T1w and T2w MRI images. We wanted to implement a defacing tool into our data ingest pipeline to make the processes of sharing neuroimaging data easier for data owners and to protect the privacy of the subjects. There a several defacing tools which have different approaches to defacing scans [16]. We investigated the suitability of two of these packages: pydeface [17] and fsl_deface [18], to see which one to implement in our data pipeline. We did this by running these two tools on T1w and T2w scans from five different cohorts held within DPUK to assess how they handle scans from multiple scanners and different qualities. This was determined through visual inspection of the scans to examine what facial features had been removed successfully and whether any important information had also been removed such as part of the brain. We also wanted to develop processes to strip any personally identifiable information (PII) from DICOM headers and JSON files if any were present in the fields.

### 4. Standardisation

Neuroimaging studies often result in thousands of files, with no standard way of structuring and naming them. Two different studies could take completely different approaches to how they structure their data and name their files, resulting in confusion when it comes to understanding these structures. It often takes time to comprehend how researchers have named their files and trying to figure out which modality a particular scan is. When it comes to automated scripts, it also makes it difficult when structures are different each time, resulting in the rewriting of scripts and rearranging of data. This isn’t very efficient and can be very time-consuming which is why Brain Imaging Data Structure (BIDS) [19] was created for datasets to follow strict ordering and naming conventions so that it is standardised across studies. Figure 1 demonstrates an example of converting an unstructured dataset into BIDS.

**Figure 1.**
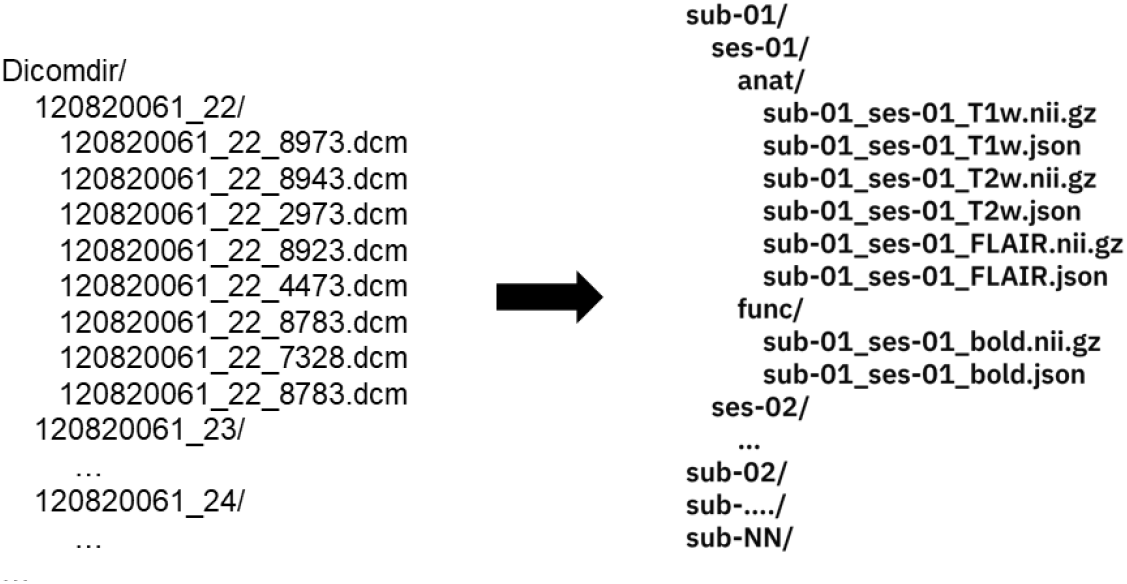
(Digital Imaging and Communications in Medicine (DICOM) to BIDS)

**Figure 2.**
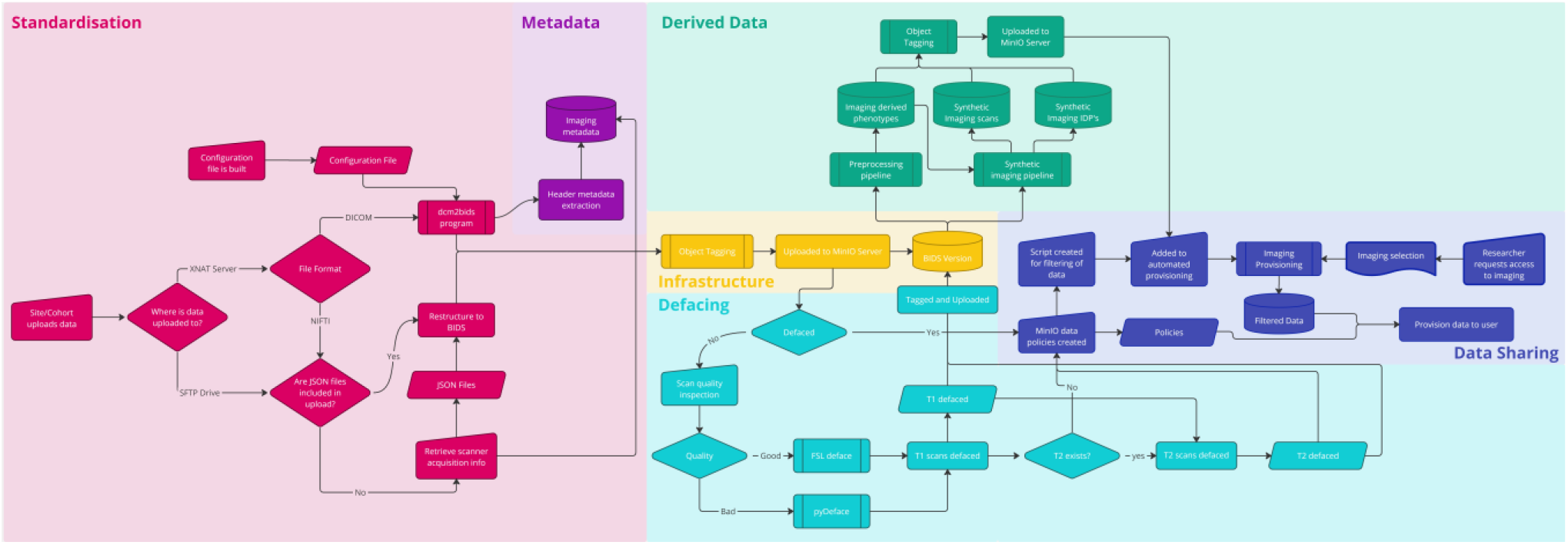
(Imaging Ingest Pipeline)

However, although this standard exists, not every study chooses to implement this structure, and instead still chooses to implement their own structure. Therefore, we decided to implement the BIDS structure for each of our imaging datasets to allow for a standardised way of structuring and naming scans to allow for automated scripts to be run across multiple datasets, allow easier understanding for the researchers to use the data and allow the data to seamlessly integrate with existing pipelines and analytical packages.

### 5. Imaging Data Sharing

Neuroimaging platforms often distribute their data through direct local downloads on to the researchers machines, whereas TRE’s will hold the data within their environment where it cannot leave. Our secure platform allows imaging data to be accessed through a secure virtual environment where that data can never leave. However, the usual file store systems available on windows is not sufficient enough to be able to store such large amounts of data in an efficient way. That’s why we have had to find solutions to effectively store data using object storage, and then give access to that data to users within the TRE. This is explored through two ways: 1) using the MinIO Python API to retrieve and work on the data and 2) mounting the data to a drive so that it displays in a file store format which people are used to. We also looked at creating an automated provisioning system to efficiently and safely provision imaging data, therefore decreasing admin time and reducing human error.

### 6. Multi-Cohort Access

DPUK operates a data governance model that enables the data provider to maintain full authority over access to the dataset. This ensures that data is only shared in accordance with the onward sharing restrictions of the data collection. This governance model has allowed over 60 data collections to be accessible via DPUK’s data portal. DPUK operates using the HDRUK five safes model to ensure that people, projects, data and computing environments are safe and secure for sharing of data [20]. This ensures that applications are processed quickly, researchers are checked, projects are assessed for scientific credibility and final decisions on access is carried out by the data provider themselves. Any results are also checked before leaving the research environment. This gives data providers’ peace of mind regarding the onward sharing of data and ensures that they are accredited for all outputs arising from use of the data. Because any cohort can deposit their data into our environment, we have had to develop automated data ingest pipelines to be able to handle the large volumes of data which come through to DPUK.

### 7. Derived Data

Processing imaging data is an essential part of most research projects to be able to extract meaningful data to perform analysis on. Researchers often perform a similar set of processing which generates a lot of extra data and therefore greater storage requirements are needed of the TRE to hold this data. This, especially, becomes a problem when there’s multiple researchers working on the same datasets, performing similar processing and therefore generating lots of additional data which has to be stored by the TRE. DPUK aims to 1) lower the storage and compute resource requirements and 2) make the process of analysing imaging data simpler by already providing derived/processed data. Users would then have the choice to choose either the raw data which they can process themselves if they wish, or to use already derived data. We plan to implement this through two different ways: one way being to ask researchers to share their derived data within the portal once they have finished their project and published results, and the other way being to implement a pre-processing pipeline within the portal to generate derived data to be requested alongside the original raw data.

## Results

We have developed a data pipeline for imaging data to meet all seven levels of imaging data facilitation criteria set out earlier in this paper. This includes several sub processes to standardise data to BIDS, extract metadata, deface scans, upload data to object storage, generate derived data, and provision data. The following sections explain these in more detail.

### 1. Metadata

In order to facilitate researchers’ understanding of the imaging data available in each cohort, an imaging matrix has been developed. This discovery tool displays available scan types and the number of participants for each cohort, enabling researchers to easily identify suitable datasets for their research and to determine which datasets can be combined to obtain the necessary data. Additionally, data insight reports are available for each cohort, providing an overview of the collected data, how the data is stored and structured, the acquisition parameters, and other relevant information that may be useful to researchers. The matrix is designed to enhance data discovery of imaging data and to simplify the process of identifying suitable cohorts for the integration of multiple imaging datasets.

Metadata was extracted from either DICOM header information or JavaScript Object Notation (JSON) sidecar files depending on the file format and were then categorised. We selected categories based on the BIDS structure so included structural MRI separated into T1w, T2w and FLAIR, functional MRI split up into rest and task, diffusion MRI, Magnetoencephalography (MEG) split up into rest and task, CT, and PET, with PET being planned to be split up further into its own distinct categories like MRI. This allows a better understanding on what data each cohort has, and also allows easier provisioning of data depending on the type of scans a researcher may want as this metadata is fed into the object tagging process.

### 2. Data & Analysis Infrastructure

When data is uploaded to DPUK, either through eXtensible Neuroimaging Archive Toolkit (XNAT) [21] or Secure File Transfer Protocol (SFTP), the data goes through our ingest pipeline and gets deposited in our MinIO object storage server. We identified MinIO as being the most suitable storage solution due to being horizontally scalable, meaning that more servers can be added easily as the storage requirements grow. This allows us to expand over time without disruptions or performance degradation and ensures that the data is evenly distributed enabling efficient utilisation of resources. There is also no vendor locking which means that we are not tied to any specific vendor therefore allowing flexibility and greater freedom. MinIO also utilises HTTPS protocol making it easier to integrate into our existing infrastructure.

The upload process primarily involves tagging objects as they are uploaded to a bucket, this means that if a 3 Tesla MRI T1w defaced scan is uploaded, then this will be tagged with: ‘MRI’, ‘T1w’, ‘Defaced’ and ‘3T’ to allow for filtering and automated provisioning of the data. This filtering system allows us to provision data without any need for duplication of data and instead only requires one master copy which can be filtered to the user. JSON policies are then generated for assigning to users to access the data, therefore allowing read only access to the imaging data. When a research project is set up, they will be provided with their own empty bucket which they can use to store their processed data and results.

The DPUK portal offers users various types of virtual desktops, both windows and Linux, depending on their requirements. The current highest windows performance desktop has 128 GB of RAM and 16 processors which makes it ideal for ML applications. The same is also available for Linux which means that processing and analysis can be performed at a faster and more efficient rate. However, for processing of large amounts of imaging data, this still often isn’t enough. That’s why DPUK has a newly launched High-Performance Computing (HPC) cluster which consists of 3 login nodes and 30 compute nodes. These run the latest AMD Epyc processors, NVMe drives and the latest server hardware giving a total of over 15T of RAM and 3500 CPU cores which can be utilised depending on the researcher’s requirements. GPU cores are now also made available which are beneficial in imaging processing. This vastly decreases the time it takes to run processing and analysis of imaging data as these tasks can be parallelised. This potentially reduces hundreds of days’ worth of processing into just a few days, meaning researchers can get started straight away with analysing the data. The HPC cluster uses singularity to install and run custom applications on the cluster. This allows researchers to run any kind of software that they require on the HPC, along with various types of processing and analysis scripts.

The DPUK portal gives users access to a multitude of neuroimaging processing and analysis tools which allow researchers to use the tools that they are most comfortable with. For the processing of neuroimaging data, there is a range of tools that researchers often use, these include: the Oxford Centre for functional MRI of the Brain (FMRIB) FSL [22], Statistical Parametric Mapping (SPM) [23], FreeSurfer [24] and the OHBA Software Library (OSL) [25]. All of these are available to use on the data portal and allow researchers to use different programming software such as Python [26], MATLAB [27] and R [28].

### 3. Anonymisation

We ran two commonly employed defacing tools, pyDeface and fsl_deface, on T1w and T2w scans obtained from five distinct cohorts employing different scanners and protocols. The primary objective was to assess the efficacy of these tools in removing facial features while preserving crucial anatomical details across scans acquired via diverse imaging methods. For the majority of scans, fsl_deface exhibited good performance in eliminating all facial features, with a few exceptions where the mouth remained visible. However, we observed instances where the tool inadvertently removed portions of the brain. Given the potential impact on the quality of scan processing outcomes, it is imperative to avoid such occurrences. Conversely, pyDeface consistently preserved the brain while occasionally failing to remove the eyes and not being able to remove the ears. Therefore, pyDeface presented a favourable balance between preserving important brain structures and removing facial features. The reasons underlying the varying defacing outcomes across different scans remain unknown. Due to incomplete scanner information for certain scans, it was challenging to establish the root cause. Consequently, we made the decision to incorporate pyDeface into our ingest pipeline as it provided a suitable trade-off.

### 4. Standardisation

There are two main ways that neuroimaging data is uploaded into DPUK: 1) through XNAT where imaging data is uploaded as DICOM or 2) SFTP where Neuroimaging Informatics Technology Initiative (NIFTI) files are uploaded directly through file transfer. As XNAT has its own standardised file structure (figure 4), this makes it slightly easier to reorganise and convert into BIDS as tools exist to be able to do this. The python package ‘dcm2bids’ [29] and ‘dcm2nii’ [30] were used to convert DICOM imaging data to NIFTI files, conforming to the BIDS file format. This package requires a configuration JSON file to map the DICOM images to BIDS which requires extracting data from DICOM headers, identifying the imaging type, and renaming the data accordingly.

**Figure 3.**
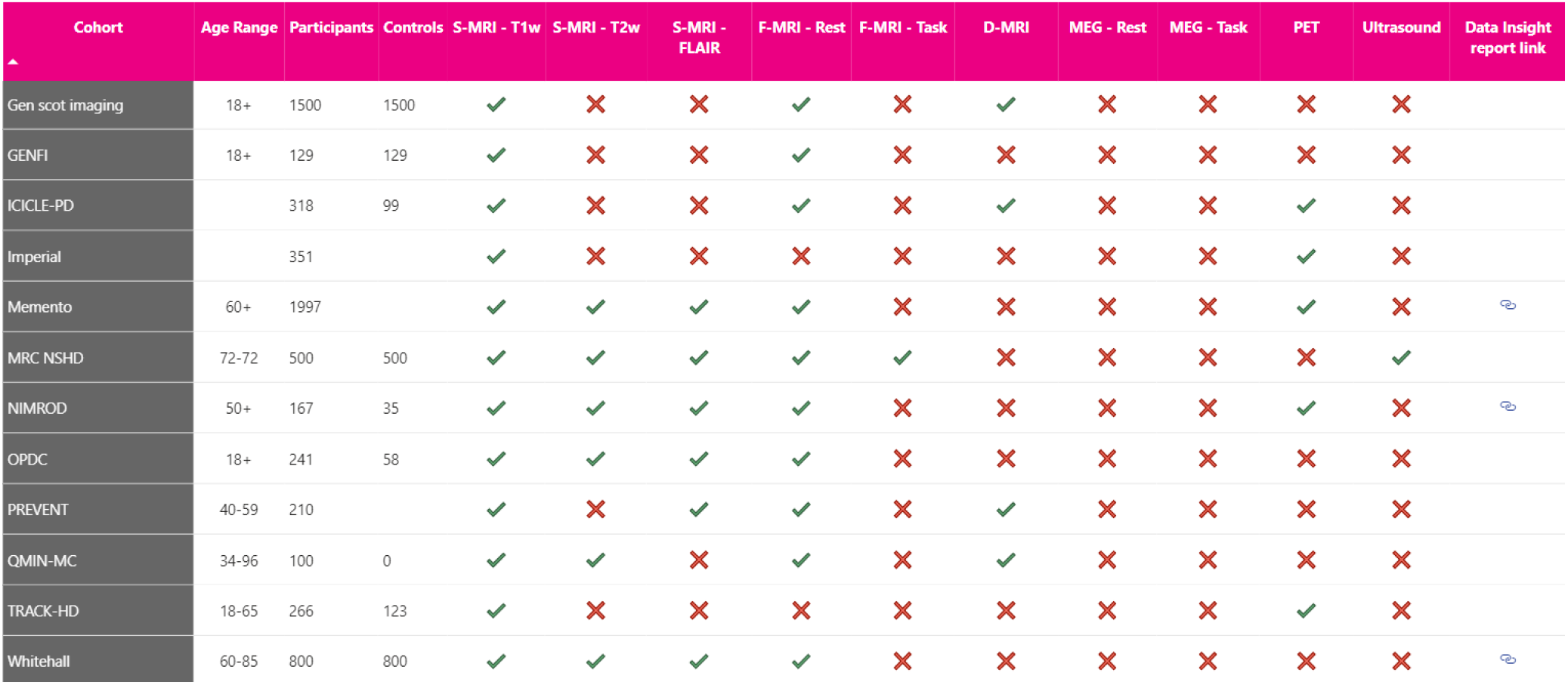
(DPUK imaging datasets – Imaging matrix of scan types)

**Figure 4.**
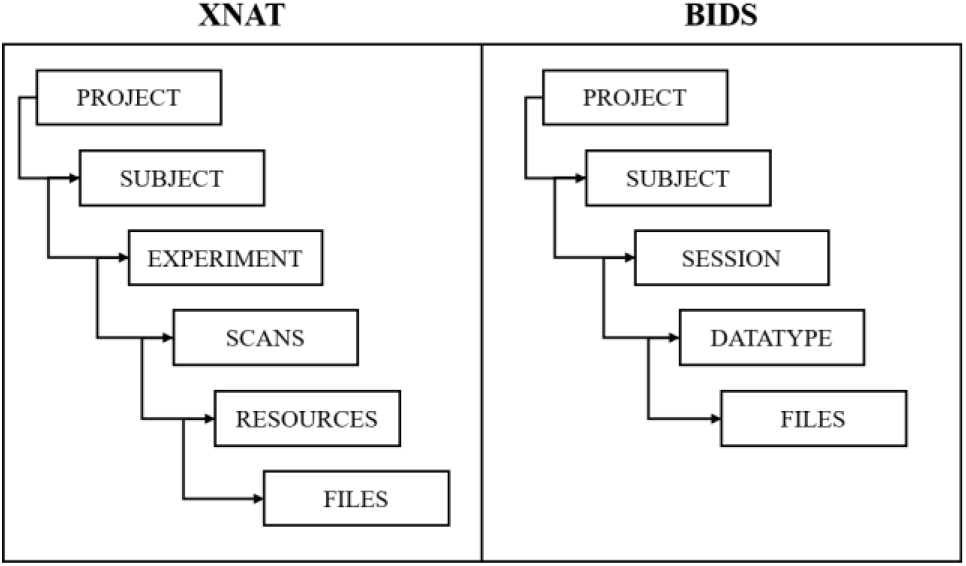
(XNAT and BIDS structures)

A semi-automated script was created to generate these configuration files from extracting data from DICOM headers. This process involves extracting the ‘Series Description’ from the DICOM header information and identifying whether certain labels are present such as if ‘t1_mprage’ was identified then this would be assigned as ‘T1w’, or if ‘diff’ was identified then this would be assigned as ‘dwi’. However, there are some instances where there are multiple variations of the same scan or sometimes the series description may not entirely match. In this case, these have to be manually inspected and assigned labels for the configuration file. Even though there is still some manual work left, this decreases the amount of time it takes to build these configuration files which can often be a lengthy process, especially if multiple centres were involved in the data acquisition. The DFP-Pilot study [31], for example, contains imaging data acquired from six different sites, all with varying ways of describing the imaging data and with multiple versions of scans.

Therefore, having a semi-automated script for generating these configuration files saves time having to go through each and every series description. When data is uploaded via SFTP, then this can come in a varying range of inconsistent file structures and naming conventions which can make it a difficult task to standardise. This means that if it isn’t already uploaded to BIDS, then a project specific script has to be created to restructure the folders and rename the files so that they conform to BIDS. This uses a similar semi-automated approach were it looks to identify certain information within the names of the files and also the folders.

As a result, DPUK has now standardised all imaging datasets currently held within our platform and we have developed several automated pipelines to deal with the wide variety of data that is uploaded. This takes away the responsibility of the data owner having to standardise, therefore allows easier onward sharing, and provides our researchers with a suitable resource to analyse data from across cohorts.

### 5. Imaging Data Sharing

Neuroimaging and genomic data are held within a MinIO server where each cohort is stored in its own separate bucket and each file stored within that bucket is stored as an object which has tags attached to it. These tags allow easy data filtering to restrict a user’s read-only access to certain data within a bucket through implementing custom policies. Policies come in the form of JSON files which contain access control rules and permissions for users and groups. For example, if a project with three users requested access to defaced T1w, T2w and rfMRI scans from cohort1 and cohort2, then a group would be set up under that project. A custom policy will then be generated through the automated provisioning program which allows users from that project group to have read-only access to any data within cohort1/cohort2 where the data has a tag of both defaced and T1w, T2w and rfMRI. This means that the rest of the data is hidden to the user and they are unable to access it. This allows just one dataset to be stored and provisioned meaning that the data is not duplicated at any time, therefore reducing storage requirements.

Users are then able to access this data on the virtual machine either through using the Python API, or by using the mounting scripts provided to the user. These mounting scripts are generated during the automated provisioning process and utilise the rclone [32] command to mount the data form the MinIO server as a drive. These mounting scripts can be run on both Windows and Linux machines to provide the user with easy access to a file structure of the data. Alternatively, the Python API can be used to retrieve and work on data and is left up to the user to decide which method to use.

### 6. Multi-Cohort Access

The DPUK portal allows the user to request and access multiple cohorts which are on the platform. This creates a centralised space where multiple datasets can be stored, accessed, processed and analysed. The DPUK portal holds around 10 neuroimaging datasets, totalling 3,735 participants, all containing multi-modal imaging data. These include structural, functional and diffusion MRI scans as well as containing PET and MEG data. This opens up possibilities which could never be achieved by using just a single dataset. The age range of these participants is from 18 to 95, and come with large phenotypic datasets, making it possible to be able to study the effect many different factors, such as lifestyle choices and diagnoses, have on the brain. Because these datasets are multi-modal, this also opens up the ability to extract many more features, allowing a great deal of opportunities to find new biomarkers for disease progression and running deeper analysis.

Because these datasets are standardised to BIDS, automated scripts can be used across multiple datasets without the need to change them based on the structure and naming of the files. This allows multiple datasets to be used to run processing and analysis without the need to rewrite scripts, and even allows the use of automated processing pipelines. This makes the initial process of preparing the data for use in machine learning applications a lot quicker for researchers, saving wasted time and effort to rewrite scripts and reorganise data. The accompanying JSON sidecars also make this process more efficient by enabling automated scripts to easily get information about the scans to be used for processing and analysis.

### 7. Derived Data

Pre-processing imaging scans is often a common essential step for researchers to carry out their analysis but can be a very time consuming and computationally expensive process which requires large amounts of storage. Because of this, we are in the process of developing and reviewing pre-processing pipelines to be implemented within our environment for researchers to use. This will eventually include a supplementary derivatives dataset which a user can apply for alongside the original data to use in their research. The derivatives data is stored in a similar format to BIDS, following it as closely as possible, so that researchers have access to a range of already processed data, therefore reducing time and resources needed. Imaging derived phenotypes (IDP’s), in the form of CSV files, will also be accessible to researchers for the cohort data they have access to. However, currently, only some of our datasets come with IDP’s provided by the data owners, or with scripts which users can run to generate derivatives. We also ask researchers, once they have published their results that they should allow their data to be shared within our environment for other researchers to use. These come as complementary datasets which researchers can select during the provisioning process. Allowing derivatives to be shared alongside the original data, significantly reduces the amount of storage required by a project and instantly gives researchers access to research-ready data.

## Discussion

### Next Steps: Pre-Processing and Synthetic Pipelines

The next stage of the ingest pipeline is to include pre-processing and synthetic data generation processes to create supplementary datasets to the original cohort data. Pre-processing pipelines are already being developed and adapted to be implemented within our environment. Once this is complete, users will be able to access harmonised derivatives and IDP’s across all imaging datasets held within DPUK. This should significantly reduce time and storage requirements for projects which choose to use this data and will remove the barrier of the often difficult process of processing and harmonisation, therefore allowing a wider range of researchers, such as those in the field of AI, to conduct analysis.

The use of synthetic data is also being explored to offer researchers with tools to generate synthetic imaging scans or synthetic IDP’s depending on demographic and disease characteristics. Several ways are being looked at for doing this including variational auto encoders (VAE’s), generative adversarial network models (GAN’s), and transformer models. Similar techniques are used within the field of AI where datasets are imbalanced or data sizes are lacking. Being able to increase the numbers of training data can considerably improve AI models and can help to reduce bias in minority groups.

## Conclusion

There are many challenges that arise with storing and analysing large neuroimaging datasets. These range from having the storage capacity to hold the data along with its subsequent derivatives, to having the performance power to run processing and analysis. With the increase of neuroimaging cohorts and research, DPUK has had to solve these problems to allow for the best experience for researchers using the data portal. This is being implemented through 1) providing metadata tools to explore imaging data available, 2) developing the data infrastructure through MinIO object storage to solve storage demands, 3) anonymising datasets by removing personally identifiable data and defacing imaging scans, 4) standardisation of all datasets to conform to BIDS, 5) imaging data sharing through our secure research environment, 6) having multi-cohort access and 7) derived data generation tools. All of this comes with a variety of tools embedded in the data portal and access to computational resources, enabling a range of techniques for researchers to employ.

## Data Availability

All data used is available through the Dementias Platform UK Data Portal at https://portal.dementiasplatform.uk/

## Acknowledgements

Funding for the DPUK Data Portal as an infrastructure support service is provided by MRC. The authors acknowledge and thank the DPUK team, without whom the work would not be possible and also the data providers that have uploaded imaging data to the DPUK data portal:

AMPLE - O’Brien, J. (2017). AMPLE [Data set]. Dementias Platform UK. https://doi.org/10.48532/005000

BioFIND - Henson, R. (2020). BioFIND [Data set]. Dementias Platform UK. https://doi.org/10.48532/007000

Cam-CAN - Henson, R. (2017). Cam-CAN [Data set]. Dementias Platform UK. https://doi.org/10.48532/009000

EPIC Norfolk - Wareham, N. (2017). EPIC Norfolk [Data set]. Dementias Platform UK. https://doi.org/10.48532/023000

Generation Scotland - Kivimaki, M. (2017). Whitehall II [Data set]. Dementias Platform UK. https://doi.org/10.48532/048000

ICICLE-PD - Burn, D. (2017). ICICLE-PD [Data set]. Dementias Platform UK. https://doi.org/10.48532/029000

Memento - Dufouil, C. (2017). MEMENTO [Data set]. Dementias Platform UK. https://doi.org/10.48532/031000

MRC NSHD - Chaturvedi, N. C. (2017). MRC NSHD [Data set]. Dementias Platform UK. https://doi.org/10.48532/035000

OPDC - Hu, M. (2017). OPDC Discovery [Data set]. Dementias Platform UK. https://doi.org/10.48532/036000

PREVENT - Ritchie, C. (2017). PREVENT [Data set]. Dementias Platform UK. https://doi.org/10.48532/038000

TRACK-HD – Tabrizi, S. (2017). TRACK HD [Data set]. Dementias Platform UK. https://doi.org/10.48532/045000

Whitehall - Kivimaki, M. (2017). Whitehall II [Data set]. Dementias Platform UK. https://doi.org/10.48532/048000

## Ethics Statement

Ethical approval was not required as only anonymised data was accessed for the purpose of this work.

## References

[1] [Internet]. Available from: https://www.who.int/publications-detail-redirect/9789240033245

[2] What are the costs of dementia care in the UK? [Internet]. Available from: https://www.alzheimers.org.uk/about-us/policy-and-influencing/dementia-scale-impact-numbers

[3] Banerjee D, Muralidharan A, Hakim Mohammed AR, Malik BH. Neuroimaging in dementia: A brief review. Cureus. 2020; doi:10.7759/cureus.8682

[4] Márquez F, Yassa MA. Neuroimaging biomarkers for alzheimer’s disease. Molecular Neurodegeneration. 2019;14(1). doi:10.1186/s13024-019-0325-5

[5] Joseph A, Jayaraman C. Preprocessing techniques for neuroimaging modalities: An in-depth analysis. Neuroimaging - New Insights [Working Title]. 2023; doi:10.5772/intechopen.109803

[6] Pellegrini E, Ballerini L, Hernandez M del, Chappell FM, González-Castro V, Anblagan D, et al. Machine learning of neuroimaging for assisted diagnosis of cognitive impairment and dementia: A systematic review. Alzheimer’s & Dementia: Diagnosis, Assessment & Disease Monitoring. 2018;10(1):519–35. doi:10.1016/j.dadm.2018.07.004

[7] Crawford KL, Neu SC, Toga AW. The image and Data Archive at the Laboratory of Neuro Imaging. NeuroImage. 2016;124:1080–3. doi:10.1016/j.neuroimage.2015.04.067

[8] Markiewicz CJ, Gorgolewski KJ, Feingold F, Blair R, Halchenko YO, Miller E, et al. The OpenNeuro resource for sharing of Neuroscience Data. eLife. 2021;10. doi:10.7554/elife.71774

[9] Gallier S, Price G, Pandya H, McCarmack G, James C, Ruane B, et al. Infrastructure and operating processes of pioneer, the HDR-UK data hub in acute care and the workings of the Data Trust Committee: A protocol paper. BMJ Health & Care Informatics. 2021;28(1). doi:10.1136/bmjhci-2020-100294

[10] Alzheimer’s Disease Data initiative (ADDI) - transforming research [Internet]. Available from: https://www.alzheimersdata.org/

[11] Davatzikos C. Machine learning in neuroimaging: Progress and challenges. NeuroImage. 2019;197:652–6. doi:10.1016/j.neuroimage.2018.10.003

[12] Fischl B. Freesurfer. NeuroImage. 2012;62(2):774–81. doi:10.1016/j.neuroimage.2012.01.021

[13] SeRP. 2022. Available from: https://serp.ac.uk/

[14] Bauermeister S, Orton C, Thompson S, Barker RA, Bauermeister JR, Ben-Shlomo Y, et al. The dementias platform UK (DPUK) Data Portal. European Journal of Epidemiology. 2020;35(6):601–11. doi:10.1007/s10654-020-00633-4

[15] MinIO Inc. High performance, kubernetes native object storage [Internet]. Available from: https://min.io/

[16] Theyers AE, Zamyadi M, O’Reilly M, Bartha R, Symons S, MacQueen GM, et al. Multisite comparison of MRI defacing software across multiple cohorts. Frontiers in Psychiatry. 2021;12. doi:10.3389/fpsyt.2021.617997

[17] Pydeface [Internet]. Available from: https://pypi.org/project/pydeface/

[18] Alfaro-Almagro F, Jenkinson M, Bangerter NK, Andersson JLR, Griffanti L, Douaud G, et al. Image processing and quality control for the first 10,000 brain imaging datasets from UK Biobank. NeuroImage. 2018;166:400–24. doi:10.1016/j.neuroimage.2017.10.034

[19] Gorgolewski KJ, Auer T, Calhoun VD, Craddock RC, Das S, Duff EP, et al. The brain imaging data structure, a format for organizing and describing outputs of neuroimaging experiments. Scientific Data. 2016;3(1). doi:10.1038/sdata.2016.44

[20] Varma S, Hubbard T, Seymour D, Brassington N, Madden S, Alliance UHDR, et al. Building Trusted Research Environments - principles and best practices; towards Tre Ecosystems [Internet]. 2021 Available from: https://zenodo.org/record/5767586

[21] Marcus DS, Olsen TR, Ramaratnam M, Buckner RL. The extensible Neuroimaging Archive Toolkit. Neuroinformatics. 2007;5(1):11–33. doi:10.1385/ni:5:1:11

[22] Jenkinson M, Beckmann CF, Behrens TEJ, Woolrich MW, Smith SM. FSL. NeuroImage. 2012;62(2):782–90. doi:10.1016/j.neuroimage.2011.09.015

[23] SPM - statistical parametric mapping [Internet]. Available from: https://www.fil.ion.ucl.ac.uk/spm/

[24] Fischl B. Freesurfer. NeuroImage. 2012;62(2):774–81. doi:10.1016/j.neuroimage.2012.01.021

[25] OSL [Internet]. Available from: https://ohba-analysis.github.io/osl-docs/#!

[26] Welcome to Python.org [Internet]. Available from: https://www.python.org/

[27] Matlab [Internet]. [cited 2023 Jul 4]. Available from: https://uk.mathworks.com/products/matlab.html

[28] The R project for statistical computing [Internet]. Available from: https://www.r-project.org/

[29] DCM2BIDS [Internet]. Available from: https://unfmontreal.github.io/Dcm2Bids/

[30] Rordenlab. Rordenlab/dcm2niix: Dcm2nii DICOM to NIfTI converter: Compiled versions available from nitrc [Internet]. Available from: https://github.com/rordenlab/dcm2niix

[31] Koychev I, Lawson J, Chessell T, Mackay C, Gunn R, Sahakian B, et al. Deep and frequent phenotyping study protocol: An observational study in prodromal alzheimer’s disease. BMJ Open. 2019;9(3). doi:10.1136/bmjopen-2018-024498

[32] Craig-Wood N. Rclone syncs your files to Cloud Storage [Internet]. Available from: https://rclone.org/

